# Reduction in risk of death among patients admitted with COVID-19 between first and second epidemic waves in New York City

**DOI:** 10.1101/2022.03.29.22273044

**Authors:** Anthony Bowen, Jason Zucker, Yanhan Shen, Simian Huang, Qiheng Yan, Medini K. Annavajhala, Anne-Catrin Uhlemann, Louise Kuhn, Magdalena Sobieszczyk, Delivette Castor

**Affiliations:** Columbia University Irving Medical Center, New York, New York, USA

**Keywords:** COVID-19, Mortality, Logistic Models

## Abstract

Many regions have experienced successive epidemic waves of COVID-19 since the emergence of SARS-CoV-2 with heterogeneous differences in mortality. Elucidating factors differentially associated with mortality between epidemic waves may inform clinical and public health strategies. We examined clinical and demographic data among patients admitted with COVID-19 during the first (March-June 2020) and second (December 2020-March 2021) epidemic waves at an academic medical center in New York City. Hospitalized patients (N=4631) had lower mortality during the second wave (14%) than the first (23%). Patients in the second wave had a lower 30-day mortality (Hazard Ratio (HR) 0.52, 95% CI 0.44, 0.61) than those in the first wave. The mortality decrease persisted after adjusting for confounders except for the volume of COVID-19 admissions (HR 0.88, 95% CI 0.70, 1.11), a measure of health system strain. Several demographic and clinical patient factors were associated with an increased risk of mortality independent of wave.

**Article summary:** Using clinical and demographic data from COVID-19 hospitalizations at a tertiary New York City medical center, we show that a reduction in mortality during the second epidemic wave was associated with decreased strain on healthcare resources.

## Introduction

By March 15, 2022, Severe Acute Respiratory Syndrome Coronavirus 2 (SARS-CoV-2), which causes Coronavirus Disease 2019 (COVID-19), has led to over 460 million confirmed infections and over 6 million deaths worldwide (*1*). New York City (NYC) experienced one of the earliest and largest local epidemics with a peak of over 16,000 daily hospitalizations and 700 daily deaths in April 2020 (*2*). COVID-19 infections in New York City declined and remained relatively low from July through November 2020 averaging fewer than 60 hospitalizations per day and 15 deaths per day during this time period (*2*). A second epidemic wave occurred from December 2020 through March 2021 resulting in a peak of nearly 400 daily hospitalizations and 90 daily deaths by February 2021(*2*).

Data from the US and Europe showed significant heterogeneity in mortality rates between the first and subsequent waves of COVID-19 (*3,4*). While many regions have reported lower case-fatality rates (CFRs) in the second epidemic wave compared to the first, some countries have demonstrated the reverse pattern (*5*–*7*). Explanations for the frequently-observed mortality reduction over time include the development and use of effective therapies, seasonal effects, viral variant effects, age, race, ethnicity, and co-morbidity differences, but these hypotheses have been under-explored (*8*). In the U.S., race, and ethnicity have been strong correlates of COVID-19 mortality and may play a role in observed differences between epidemic waves (*9*). Among regions with a trend toward increased mortality in the second wave, proposed explanations include increased pressure on the healthcare system as well as the emergence of new viral variants (*7*). Previous studies reporting CFRs between epidemic periods were not able to examine the impact of related demographic, health system, or environmental factors.

Here, we investigated whether mortality differed by epidemic wave; and whether individual-level demographic (e.g., age, race, and ethnicity) and clinical factors, as well as markers of health system burden, affected mortality among COVID-19 patients admitted to an academic medical center and an affiliated community hospital in New York City.

## Methods

### Data Sources

The study was conducted at a large quaternary academic medical center and an affiliated community hospital in New York, NY, USA. Patients included were admitted with a positive or presumed positive SARS-CoV-2 RT-PCR test within two days of hospital presentation between March 1, 2020 and March 31, 2021. Data were extracted and cleaned from the medical center clinical data warehouse and electronic health record (EHR) as previously described (*10*–*12*). Patient demographics, anthropometric measurements, SARS-CoV-2 RT-PCR cycle threshold (Ct) value, level of respiratory support, Intensive Care Unit (ICU) admission status, historical and current medications, and discharge status were collected. Study approvals were obtained from the Columbia University Irving Medical Center Institutional Review Board (IRB), New York, NY, USA. The requirement for obtaining written informed consent was waived by the IRB.

### Variables Assessed

We defined COVID-19 cases in our cohort by three epidemic periods; the interval where cases increased, peaked, and decreased, were called waves. The first wave was defined from March 1, 2020, to June 30, 2020; the inter-epidemic period from July 1, 2020, to November 30, 2020; and the second wave from December 1, 2020, to March 31, 2021. Sex, age, race, and ethnicity were self-reported. Body mass index (BMI) was collected on admission and calculated by dividing weight in kilograms by the square of height in meters. BMI was categorized using a ≥ 30kg/m^2^ cut point for obese and < 30kg/m^2^ as normal. Viral load assessments based on SARS-CoV-2 RT-PCR Ct values were reported for cobas (Roche Molecular Systems, Inc., Branchburg, NJ), and Xpert Xpress assays (Cepheid, Inc., Sunnyvale, CA), but not for the BioFire Respiratory Panel assay (BioFire Diagnostics, Salt Lake City, UT). The ORF1ab gene was targeted for the cobas assay and the N2 gene was targeted for the Xpert Xpress assays.

Quantitative Ct values were converted to high, medium, and low viral load categories based on tertiles. For the cobas assay, high, medium, and low viral load was defined by Ct values <25, 25-30, and >30, respectively. For the Xpert Xpress assay, high, medium, and low viral load were defined by Ct values <27, 27-32, and >32, respectively. Choice of viral load assay was based on laboratory needs, resources, and timing. The level of respiratory support at hospital presentation was recorded as: room air, nasal cannula, non-rebreather, non-invasive ventilation, or intubation. We also recorded whether patients or their decision-makers elected for Do-Not-Intubate (DNI) status. Patients admitted into an ICU within 24 hours of hospital admission were considered as admitted to an ICU at presentation. Steroid usage was defined by documented receipt of intravenous or oral formulations of prednisone, dexamethasone, or methylprednisolone. Use of remdesivir, the first antiviral agent approved by the Food and Drug Administration (FDA) for COVID-19, was also recorded (*13*). Underlying coronary artery disease (CAD), chronic kidney disease (CKD), diabetes mellitus (DM), or hypertension (HTN) was defined by current or historical International Classification of Disease (ICD-10) codes (Table S1). We calculated the age-adjusted Charlson Comorbidity Index (CCI) using the EHR (*14*). Hospital Frailty Risk Score (HFRS) was calculated among patients ≥ 75 years old (*15*). The weekly number of SARS-CoV-2 admissions were recorded as a proxy of the hospital COVID-19 burden. The primary outcome was death or discharge to hospice. Survival time was calculated as days from the date of hospital admission to the date of death or discharge to hospice for events and from admission to discharge alive for the rest.

### Statistical Analyses

Histogram plots were used to visualize the distribution of COVID-19 cases and admissions. Descriptive statistics were reported including counts with percentages, medians and their interquartile ranges (IQRs), and box-and-whisker plots. The Wilcoxon rank-sum test was used to compare groups for continuous variables, and χ□^2^ test was used for categorical variables. Unadjusted logistic regression analyses were used to estimate the associations between epidemic wave and patient demographic, anthropometric, clinical, and viral load characteristics. The wave was defined as a binary variable and the inter-epidemic period was excluded in regression analyses.

Mortality was examined using Kaplan-Meier survival analysis and Cox Proportional Hazards models. Deaths included those discharged to hospice and time to death was calculated from the date of admission. Those who did not die were considered alive until March 31, 2021. In Cox Proportional Hazards analyses, survival times were right censored on day 30 after admission. Proportional hazards assumption was examined through graphical examination. Final models focused on 30-day survival and investigated potential covariables in conjunction with the epidemic wave. Models were also run separately for those ≥ 75 years old. All statistical analyses were conducted using R Studio (Boston, MA, USA).

## Results

Patients who met inclusion criteria (N=4631) were grouped by date of admission into the first wave (March 1, 2020, to June 30, 2020; N=2846), an inter-epidemic period (July 1, 2020, to November 30, 2020; N=366), and the second wave (December 1, 2020, to March 31, 2021; N=1419).

The volume of SARS-CoV-2 cases and admissions (Figure 1A-B) vastly differed between waves 1 and 2. The median length of hospitalization among patients who died or were discharged to hospice (Figure 1C) was shorter in wave 1 than 2. The distribution of length of hospital stay among patients who were discharged alive did not differ by epidemic wave (Figure 1D). Monthly mortality rate (per 100 inpatients) peaked at >25% in April 2020 during the first wave, declined to 5-10% during the inter-epidemic period, and rose to 15% during the second wave (Figure S1).

**Figure 1.**
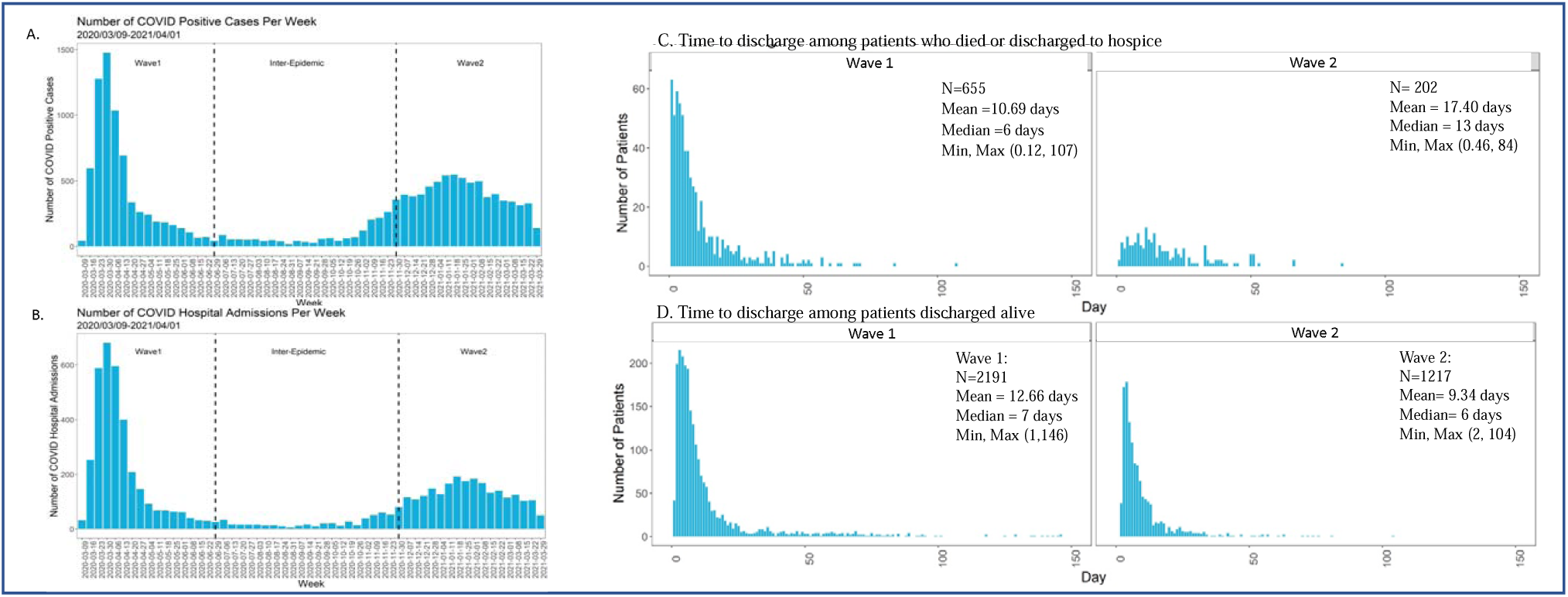
Distribution of all SARS-Cov-2 cases (A), SARS-CoV-2 admissions (B), time to death among patients who died or were discharged to hospice (C), and hospital length of stay among patients who were discharged alive (D).

Table 1 shows patient characteristics by epidemic period and the unadjusted association between each covariate and wave. Wave 2 patients were 0.56 (95% CI 0.47, 0.66) times as likely to experience death, representing 23% of wave 1 patients and 14% of wave 2 patients. Patients in the inter-epidemic period were 0.36 (95% CI 0.26, 0.52) times as likely to experience death as those in wave 1. Age, history of co-morbidities, Charlson comorbidity index, and hospital frailty risk score did not differ by wave. Patients during the second wave were less likely to identify as male (51% versus 57%), identify as Non-Hispanic Black (11% versus 14%), to have DNI status (25% versus 33%), and be admitted to the ICU at presentation (7% versus 11%). Wave 2 patients were 1.31 (95% CI 1.02, 1.69) times as likely to have a low Ct value (high viral load). However, only 23% of patients in the second wave had recorded Ct values compared with 97% during the first wave due to the use of different testing assays. Patients during the second wave were less likely to require supplemental oxygen (58% versus 65%), non-rebreather mask (5% versus 22%), and invasive mechanical ventilation (4% versus 6%) at presentation. Patients in the second wave were also more likely to receive supplemental oxygen via nasal cannula (48% versus 36%) and non-invasive ventilation (2% versus 1%). Steroid and remdesivir use in wave 2 were 7.04 (95% CI 6.12, 8.10) and 24.77-fold (95% CI 19.41, 31.60) higher than wave 1, respectively. Weekly COVID-19 admissions divided by 50 were 0.54 times less in wave 2 compared to wave 1 (95% CI 0.52, 0.57).

**Table 1:** Demographic and clinical characteristics of patients hospitalized with COVID-19 in the first wave, second wave, and inter-epidemic period.

|  | N | Overall<br>N=4631 | First Wave*<br>N=2846 | Inter-epidemic*<br>N=366 | Second<br>wave*<br>N=1419 | OR (95% CI)** |
| --- | --- | --- | --- | --- | --- | --- |
| Discharge status, N (%) | 4631 |  |  |  |  |  |
| Alive |  | 3738 (81%) | 2191 (77%) | 330 (90%) | 1217 (86%) | Ref |
| Death/Hospice |  | 893 (19%) | 655 (23%) | 36 (10%) | 202 (14%) | **0.56 (0.47, 0.66)** |
| Status at 30 days, N (%) | 4631 |  |  |  |  |  |
| Alive or discharged |  | 3835 (83%) | 2246 (79%) | 338 (92%) | 1251 (88%) | Ref |
| Death/Hospice |  | 796 (17%) | 600 (21%) | 28 (8%) | 168 (12%) | **0.50 (0.42, 0.60)** |
| Age (years), mean (SD) | 4631 | 65.5 (17.4) | 65.4 (17.1) | 63.0 (18.5) | 66.1 (17.6) | 1.00 (1.00, 1.01) |
| Age (years), N (%) | 4631 |  |  |  |  |  |
| 18-50 |  | 869 (19%) | 517 (18%) | 87 (24%) | 265 (19%) | Ref |
| 50-65 |  | 1208 (26%) | 776 (27%) | 94 (26%) | 338 (24%) | 0.85 (0.70, 1.03) |
| 65-75 |  | 1060 (23%) | 662 (23%) | 86 (23%) | 312 (22%) | 0.92 (0.75, 1.12) |
| 75+ |  | 1494 (32%) | 891 (31%) | 99 (27%) | 504 (36%) | 1.10 (0.92, 1.33) |
| Sex, N (%) | 4631 |  |  |  |  |  |
| Female |  | 2101 (45%) | 1235 (43%) | 166 (45%) | 700 (49%) | Ref |
| Male |  | 2530 (55%) | 1611 (57%) | 200 (55%) | 719 (51%) | **0.79 (0.69, 0.89)** |
| Race/Ethnicity, N (%) | 4631 |  |  |  |  |  |
| Hispanic/Latino |  | 2398 (52%) | 1474 (52%) | 197 (54%) | 727 (51%) | Ref |
| Non-Hispanic Black |  | 586 (13%) | 400 (14%) | 32 (9%) | 154 (11%) | **0.78 (0.63, 0.96)** |
| Non-Hispanic White |  | 536 (12%) | 308 (11%) | 48 (13%) | 180 (13%) | 1.18 (0.96, 1.45) |
| Other |  | 1111 (24%) | 664 (23%) | 89 (24%) | 358 (25%) | 1.09 (0.93, 1.28) |
| BMI (kg/m2), N (%) | 4301 |  |  |  |  |  |
| <30 |  | 2766 (64%) | 1666 (64%) | 233 (67%) | 867 (63%) | Ref |
| ≥30 |  | 1535 (36%) | 920 (36%) | 113 (33%) | 502 (37%) | 1.05 (0.91, 1.20) |
| Ct value, median (IQR) | 3324 | 28 (23, 33) | 29 (24, 33) | 26 (17, 33) | 27 (22, 33) | 0.99 (0.97, 1.00) |
| Viral load categories***, N (%) | 3319 |  |  |  |  |  |
| Low (Ct >32 or 30) |  | 1400 (42%) | 1195 (43%) | 77 (33%) | 128 (39%) | Ref |
| Medium (Ct 27-32 or 25-30) |  | 778 (23%) | 667 (24%) | 38 (16%) | 73 (22%) | 1.02 (0.75, 1.38) |
| High (Ct <27 or 25) |  | 1141 (34%) | 893 (32%) | 120 (51%) | 128 (39%) | **1.34 (1.03, 1.74)** |
| Ever DNI, N (%) | 4631 |  |  |  |  |  |
| Yes |  | 1344 (29%) | 941 (33%) | 53 (14%) | 350 (25%) | **0.66 (0.57, 0.76)** |
| No |  | 3287 (71%) | 1905 (67%) | 313 (86%) | 1069 (75%) | Ref |
| Oxygen level at presentation, N (%) | 4631 |  |  |  |  |  |
| Room Air |  | 1816 (39%) | 998 (35%) | 219 (60%) | 599 (42%) | Ref |
| Nasal Cannula |  | 1835 (40%) | 1036 (36%) | 123 (34%) | 676 (48%) | 1.09 (0.94, 1.25) |
| Non-rebreather |  | 699 (15%) | 622 (22%) | 9 (2%) | 68 (5%) | **0.18 (0.14, 0.24)** |
| Non-invasive ventilation |  | 50 (1.1%) | 19 (1%) | 7 (2%) | 24 (2%) | **2.10 (1.15, 3.92)** |
| Intubation |  | 231 (5.0%) | 171 (6%) | 8 (2%) | 52 (4%) | **0.51 (0.36, 0.70)** |
| ICU admission by time, N (%) | 4631 |  |  |  |  |  |
| Non-ICU |  | 3769 (81%) | 2262 (79%) | 302 (83%) | 1205 (85%) | Ref |
| ICU at presentation |  | 437 (10%) | 309 (11%) | 34 (9%) | 94 (7%) | **0.57 (0.45, 0.72)** |
| ICU after presentation |  | 425 (9%) | 275 (10%) | 30 (8%) | 120 (8%) | 0.82 (0.65, 1.02) |
| Steroid use, N (%) | 4501 |  |  |  |  |  |
| Yes |  | 1913 (43%) | 712 (26%) | 191 (53%) | 1010 (72%) | **7.32 (6.34, 8.46)** |
| No |  | 2588 (57%) | 2026 (74%) | 169 (47%) | 393 (28%) | Ref |
| Remdesivir use, N (%) | 4631 |  |  |  |  |  |
| Yes |  | 817 (18%) | 82 (3%) | 134 (37%) | 601 (42%) | **24.8 (19.5, 31.8)** |
| No |  | 3814 (82%) | 2764 (97%) | 232 (63%) | 818 (58%) | Ref |
| History of Coronary Artery Disease, N (%) | 4631 | 730 (16%) | 433 (15%) | 65 (18%) | 232 (16%) | 1.09 (0.91, 1.29) |
| History of Chronic Kidney Disease, N (%) | 4631 | 798 (17%) | 484 (17%) | 64 (17%) | 250 (18%) | 1.04 (0.88, 1.23) |
| History of Diabetes, N (%) | 4631 | 1824 (39%) | 1139 (40%) | 148 (40%) | 537 (38%) | 0.91 (0.80, 1.04) |
| History of Hypertension, N (%) | 4631 | 2741 (59%) | 1689 (59%) | 217 (59%) | 835 (59%) | 0.98 (0.86, 1.12) |
| Age adjusted Charlson comorbidity score, median (IQR) | 4582 | 4 (2, 5) | 4 (2, 5) | 3 (1, 5) | 4 (2, 5) | 1.02 (0.99, 1.04) |
| Age adjusted Charlson comorbidity | 4582 |  |  |  |  |  |
| index, N (%) |  |  |  |  |  |  |
| 0-1 |  | 931 (20%) | 546 (19%) | 94 (26%) | 291 (21%) | Ref |
| 2-3 |  | 1318 (29%) | 838 (30%) | 93 (26%) | 387 (27%) | 0.87 (0.72, 1.04) |
| 4-5 |  | 1358 (30%) | 843 (30%) | 107 (29%) | 408 (29%) | 0.91 (0.75, 1.09) |
| 6+ |  | 975 (21%) | 581 (21%) | 70 (19%) | 324 (23%) | 1.05 (0.86, 1.27) |
| Hospital Frailty Risk Score among age ≥ 75, mean (SD) | 1476 | 6.3 (5.9) | 6.4 (6) | 7.1 (6.3) | 6.0 (5.5) | 0.99 (0.97, 1.01) |
| SARS-Cov-2 hospital admission volume per week, mean (SD) | 4631 | 320 (245) | 458 (218) | 31 (17) | 119 (34) | **0.99 (0.98, 0.99)** |
| SARS-Cov-2 hospital admission volume per week divided by 50, mean (SD) | 4631 | 6.6 (4.8) | 9.3 (4.3) | 0.7 (0.4) | 2.9 (0.6) | **0.54 (0.52, 0.57)** |

\*Epidemic waves were defined as: First wave, March 1, 2020 to June 30, 2020; Inter-epidemic, July 1, 2020 to November 30, 2020; Second wave, December 1, 2020 to March 31, 2021. \*\*The odds ratio (OR) of the characteristic in the second wave relative to the first wave. \*\*\*Viral load categories: For the cobas assay, high, medium and low viral load was defined by Ct <25, 25-30, and >30, respectively. For the Xpert Xpress assay, high, medium and low viral load was defined by Ct <27, 27-32, >32, respectively.

Figure 2A-D show Kaplan-Meier plots comparing survival between wave 1, inter-epidemic period, and wave 2 (log-rank test, p<0.001). For wave 1, the cumulative survival probabilities declined from 0.87 on day 7 to 0.79 by day 30 after admission, worse than in wave 2 where these probabilities were 0.97 at day 7 and 0.88 by day 30 (Figure 2). Survival probabilities were lower among patients ≥ 75 years old across both waves but the pattern of improved survival in wave 2 persisted (Figure 2).

**Figure 2.**
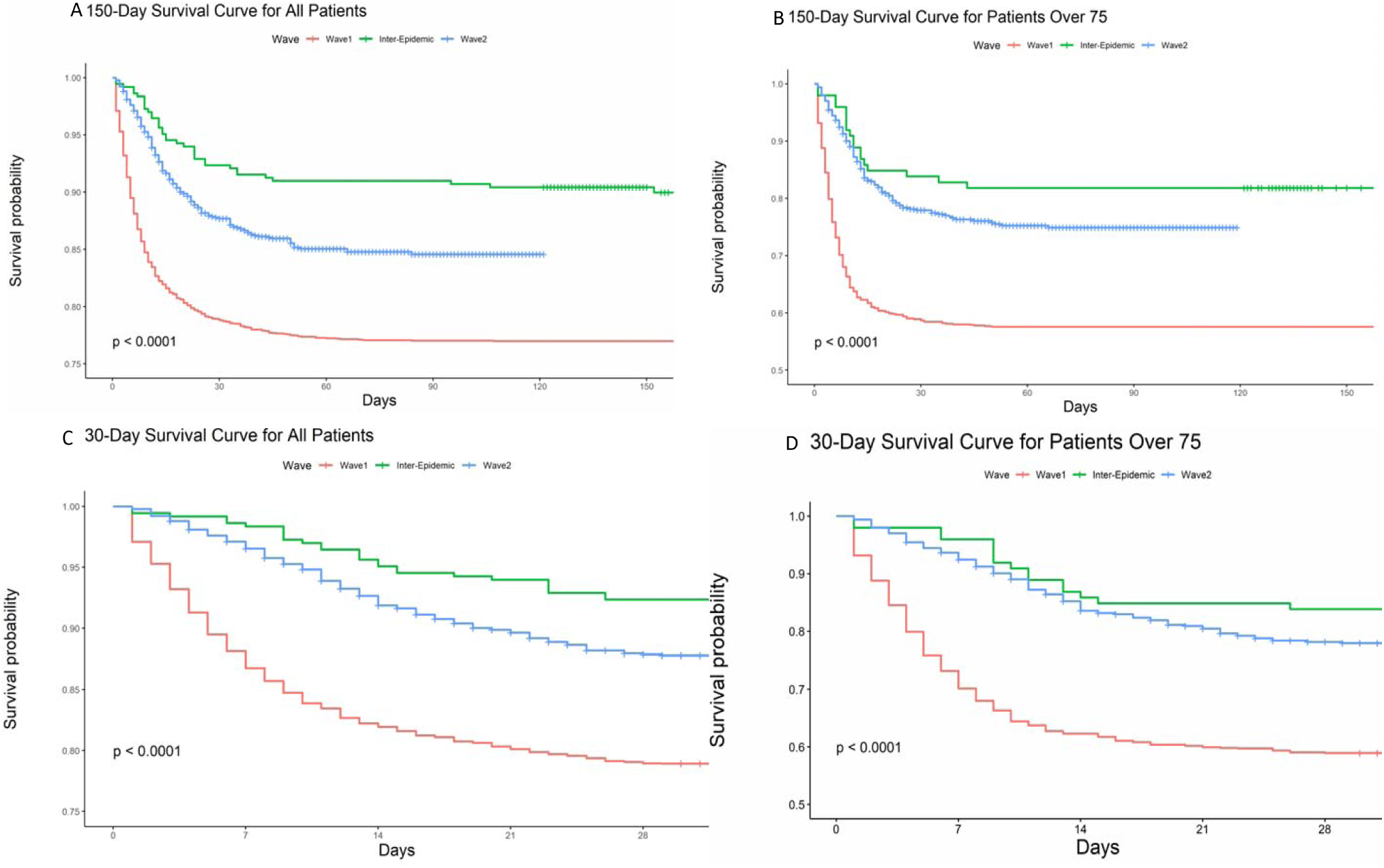
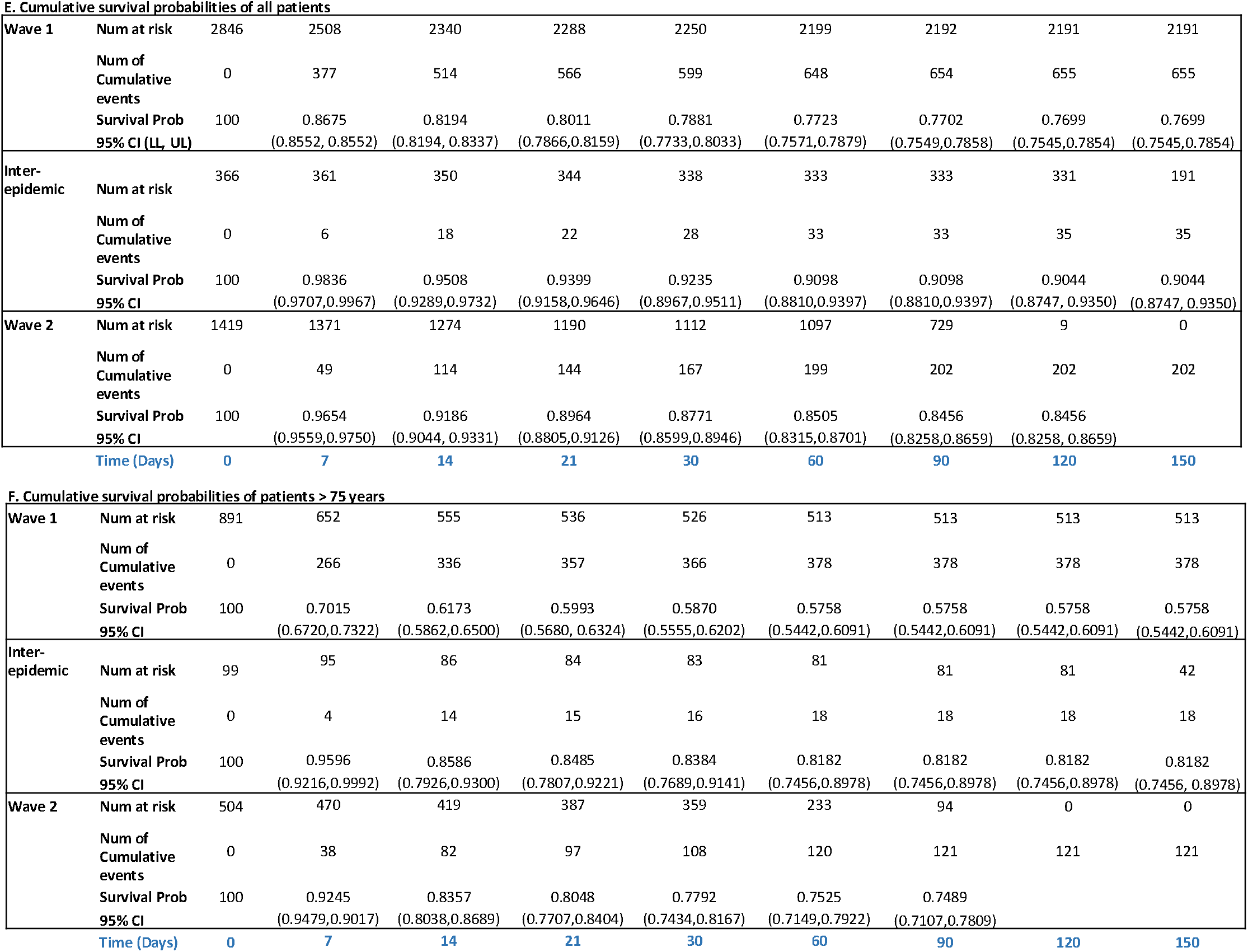
Kaplan-Meier survival plots of all patients (A) and patients ≥ 75 years old (B) hospitalized in the first and second wave of COVID-19 in New York City censored on March 31, 2021, all patients (C) and patients ≥ 75 years old (D) censored at 30 days, and cumulative survival probabilities for all patients (E), and patients ≥ 75 years old (F).

Unadjusted Cox regression for 30-day survival showed a 0.52-fold (95% CI 0.44, 0.61) reduction in risk of death in wave 2 compared to wave 1 (Table 2). The lower risk of death associated with wave 2 persisted after adjusting for potential demographic confounders. For example, after adjusting for age, sex, and race individually, wave 2 was associated with 0.46 (95% CI 0.38, 0.54), 0.52 (0.44, 0.61), and 0.51-fold (0.43, 0.61) lower mortality rate, respectively than wave 1. Oxygen level at presentation, a marker of the severity of disease, attenuated the association between wave and mortality, although hazard ratios remained below 1 and statistically significant. After adjusting for the volume of weekly COVID-19 admissions, a marker of health service strain, wave 2 was no longer associated with lower mortality (HR=0.88, 95% CI 0.70, 1.11).

**Table 2:**
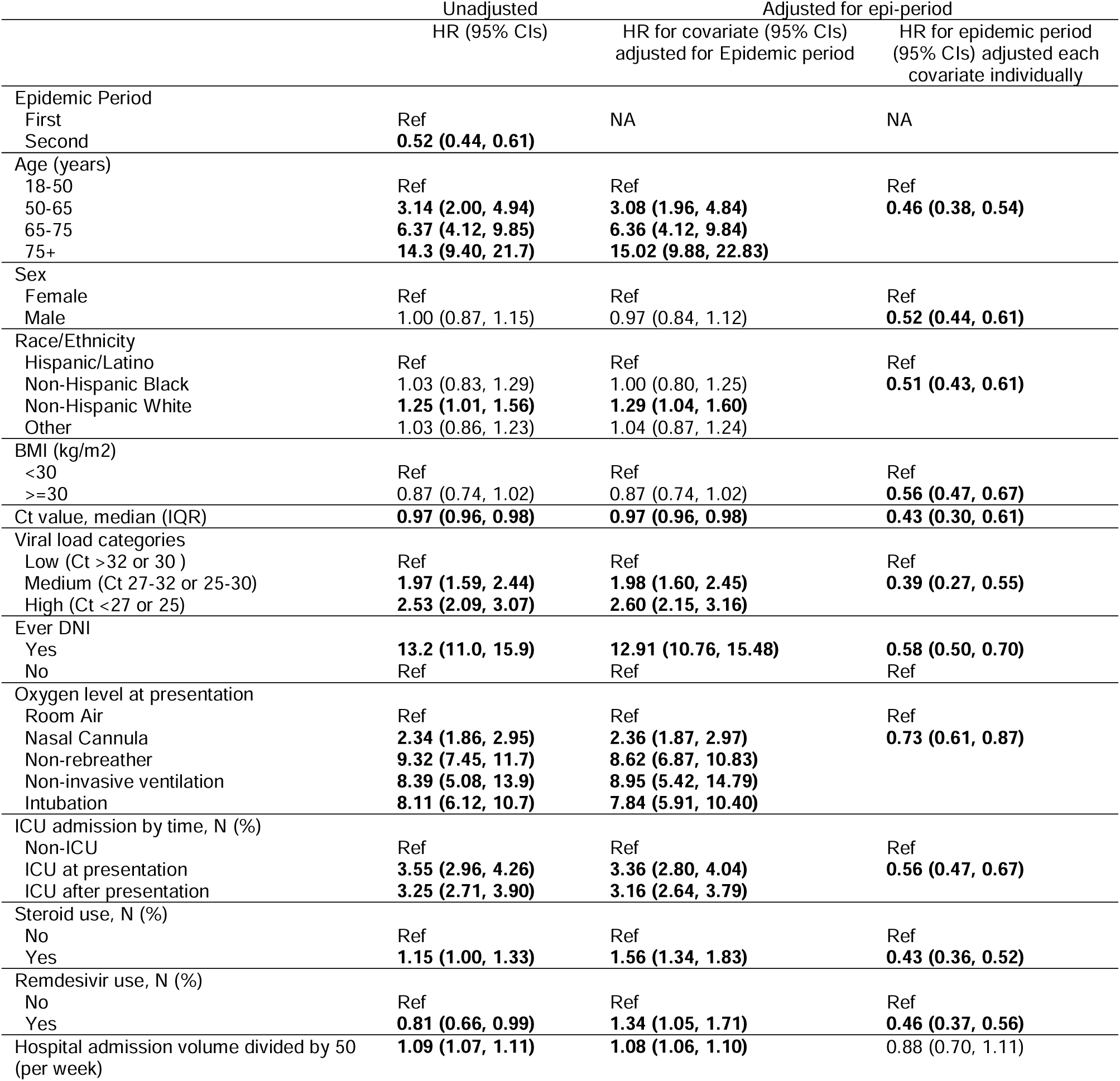
Unadjusted and adjusted Cox proportional hazards model of the association between epidemic wave and death by 30 days after admission adjusted for potentially confounding factors, among all patients.

|  | Unadjusted | Adjusted for epi-period |  |
| --- | --- | --- | --- |
|  | HR (95% CIs) | HR for covariate (95% CIs)<br>adjusted for Epidemic period | HR for epidemic period<br>(95% CIs) adjusted each<br>covariate individually |
| Epidemic Period |  |  |  |
| First | Ref | NA | NA |
| Second | **0.52 (0.44, 0.61)** |  |  |
| Age (years) |  |  |  |
| 18-50 | Ref | Ref | Ref |
| 50-65 | **3.14 (2.00, 4.94)** | **3.08 (1.96, 4.84)** | **0.46 (0.38, 0.54)** |
| 65-75 | **6.37 (4.12, 9.85)** | **6.36 (4.12, 9.84)** |  |
| 75+ | **14.3 (9.40, 21.7)** | **15.02 (9.88, 22.83)** |  |
| Sex |  |  |  |
| Female | Ref | Ref | Ref |
| Male | 1.00 (0.87, 1.15) | 0.97 (0.84, 1.12) | **0.52 (0.44, 0.61)** |
| Race/Ethnicity |  |  |  |
| Hispanic/Latino | Ref | Ref | Ref |
| Non-Hispanic Black | 1.03 (0.83, 1.29) | 1.00 (0.80, 1.25) | **0.51 (0.43, 0.61)** |
| Non-Hispanic White | **1.25 (1.01, 1.56)** | **1.29 (1.04, 1.60)** |  |
| Other | 1.03 (0.86, 1.23) | 1.04 (0.87, 1.24) |  |
| BMI (kg/m2) |  |  |  |
| <30 | Ref | Ref | Ref |
| >=30 | 0.87 (0.74, 1.02) | 0.87 (0.74, 1.02) | **0.56 (0.47, 0.67)** |
| Ct value, median (IQR) | **0.97 (0.96, 0.98)** | **0.97 (0.96, 0.98)** | **0.43 (0.30, 0.61)** |
| Viral load categories |  |  |  |
| Low (Ct >32 or 30) | Ref | Ref | Ref |
| Medium (Ct 27-32 or 25-30) | **1.97 (1.59, 2.44)** | **1.98 (1.60, 2.45)** | **0.39 (0.27, 0.55)** |
| High (Ct <27 or 25) | **2.53 (2.09, 3.07)** | **2.60 (2.15, 3.16)** |  |
| Ever DNI |  |  |  |
| Yes | **13.2 (11.0, 15.9)** | **12.91 (10.76, 15.48)** | **0.58 (0.50, 0.70)** |
| No | Ref | Ref | Ref |
| Oxygen level at presentation |  |  |  |
| Room Air | Ref | Ref | Ref |
| Nasal Cannula | **2.34 (1.86, 2.95)** | **2.36 (1.87, 2.97)** | **0.73 (0.61, 0.87)** |
| Non-rebreather | **9.32 (7.45, 11.7)** | **8.62 (6.87, 10.83)** |  |
| Non-invasive ventilation | **8.39 (5.08, 13.9)** | **8.95 (5.42, 14.79)** |  |
| Intubation | **8.11 (6.12, 10.7)** | **7.84 (5.91, 10.40)** |  |
| ICU admission by time, N (%) |  |  |  |
| Non-ICU | Ref | Ref | Ref |
| ICU at presentation | **3.55 (2.96, 4.26)** | **3.36 (2.80, 4.04)** | **0.56 (0.47, 0.67)** |
| ICU after presentation | **3.25 (2.71, 3.90)** | **3.16 (2.64, 3.79)** |  |
| Steroid use, N (%) |  |  |  |
| No | Ref | Ref | Ref |
| Yes | **1.15 (1.00, 1.33)** | **1.56 (1.34, 1.83)** | **0.43 (0.36, 0.52)** |
| Remdesivir use, N (%) |  |  |  |
| No | Ref | Ref | Ref |
| Yes | **0.81 (0.66, 0.99)** | **1.34 (1.05, 1.71)** | **0.46 (0.37, 0.56)** |
| Hospital admission volume divided by 50<br>(per week) | **1.09 (1.07, 1.11)** | **1.08 (1.06, 1.10)** | 0.88 (0.70, 1.11) |

Table 2 also shows increasing age, identifying as non-Hispanic white, lower Ct values, DNI status, supplementary oxygen requirement at presentation, ICU admission at presentation, and the volume of COVID-19 admissions were each associated with higher mortality rate after adjusting for wave. Among patients ≥ 75 years of age, wave 2 had a 0.43-fold (95% CI 0.35, 0.53) reduced risk of death compared to wave 1 (Table S2), and the mortality pattern was similar to that seen in the overall sample.

We expected steroid and remdesivir use to be associated with reduced mortality but recognize that confounding by indication may produce results showing the opposite. Therefore, we conducted stratified analyses by wave and ICU status (Table S3). These analyses suggested that steroid and remdesivir effects were modified by wave i.e., lowered mortality risk in wave one, ICU patients, and no benefit or slightly increased mortality risk in wave two. Particularly in wave 1, there was a strong suggestion of confounding by indication for steroids.

## Discussion

NYC, like many other parts of the world, has experienced multiple distinct epidemic waves of COVID-19 (*2*–*5,7*). Our analysis of 4631 patients admitted with SARS-CoV-2 during the first two epidemic waves and the inter-epidemic period revealed a decrease in risk of death or discharge to hospice in wave 2 compared to the wave 1. The association between wave and mortality persisted after covariate adjustment for several factors including age, sex, race, and markers of disease severity. However, the association between wave and mortality disappeared after adjusting for the volume of COVID-19 admissions suggesting that strain on hospital resources may have been one of the factors accounting for the high mortality rate in epidemic wave 1. Although the duration of hospital stay did not differ among patients who were discharged alive, the median time to death in the second wave was nearly one week longer than in wave 1.

There were several other variables correlated with decreased mortality. Patients presenting in the second wave were less likely to require oxygen at presentation. Among those who did require oxygen, patients in the second wave were more likely to require nasal cannula, suggesting that their disease was less severe at the time of presentation. It is likely that during the first wave, when hospitals in NYC were widely reported to be overwhelmed, patients may have been more reluctant to present to the hospital until they developed a greater degree of respiratory distress, resulting in a higher chance of intubation on arrival. We did not observe differences in frailty among patients age ≥ 75 years old, individual co-morbidities or Charlson comorbidity index over time.

Interventions may also account for differences in the mortality between the two waves. Non-invasive ventilation was commonly avoided early in the pandemic due to concerns about aerosolizing the virus as well as the theory that early intubation would lead to less risk of lung injury (*16,17*). This approach was later questioned, and subsequent studies demonstrated no benefit to early intubation for COVID-19, leading to increased use of non-invasive ventilation during the second epidemic wave (*16*–*18*). We observed effect modification by epidemic wave, and a paradoxical effect of COVID-19 therapies on mortality in wave two. Therapeutic use of low-dose corticosteroid and remdesivir were associated with lower mortality in wave 1. We used ICU status as a proxy for disease severity and it partially explained the association between steroid and remdesivir use and increased mortality observed in wave 2. Early in the COVID-19 pandemic, corticosteroids were proposed as a potential intervention to counteract progression to acute respiratory distress syndrome (ARDS). Still, their use was not routine in many centers in part due to a lack of supportive data for corticosteroids in ARDS due to influenza (*19*). In July 2020, the RECOVERY group published data showing a reduction in mortality with the use of dexamethasone among patients receiving supplemental oxygen, resulting in widespread adoption of corticosteroid use for patients admitted with COVID-19 (*20*). We observed a much higher rate of steroid use among patients in the second wave, which may have contributed to that group’s lower overall mortality rate, and associated with mortality due to the residual confounding effect of use by disease severity that we could not measure or control for in this analysis. It is likely that other changes in the clinical management of COVID-19 patients based on accumulated data throughout the pandemic similarly contributed to the lower mortality. Remdesivir became a widely used anti-viral for admitted patients requiring oxygen and the first anti-viral to be FDA-approved for COVID-19. However, it was only shown to shorten time to recovery rather than reduce mortality (*13,21*). Monoclonal antibody therapies also became available for patients early in the course of infection with mild symptoms (*22,23*). Early proning of patients requiring respiratory support was also associated with improved ventilation and outcomes (*24,25*). As data were gathered throughout the early months of the pandemic, clinicians and hospitals rapidly assembled and distributed COVID-19 management guidelines to delineate the most evidence-based and proven interventions. These protocols undoubtedly led to better uniformity in clinical practice, decreased use of unproven or ineffective therapies, and greater use of treatments with the potential to reduce mortality.

In December 2020, the FDA granted emergency use authorization (EUA) to mRNA-based COVID-19 vaccines developed by Pfizer/BioNTech and Moderna (*26*–*29*). Increasing prevalence of vaccination during the second epidemic wave in NYC may have contributed to decreases in COVID-19 admissions, especially among high-risk groups. Baseline patient characteristics including age, sex, race/ethnicity, BMI, and the presence of several co-morbidities were similar between the two epidemic waves, suggesting that the availability of vaccines did not alter the overall demographics of patients admitted with COVID-19 through March 31, 2021. We suspect that vaccination had a limited impact on mortality in the second wave since vaccine uptake in the population at risk by March 31, 2021 was still highly limited.

Rapid increases in COVID-19 cases during epidemic waves put substantial pressure on healthcare systems worldwide. During the first wave in NYC, many hospitals were overwhelmed with the rapid influx of patients combined with staff and equipment shortages, including limited ventilators, personal protective equipment (PPE), and certain essential medications. Studies have shown the critical importance of adequate medical resources with COVID-19 mortality inversely-correlated with available hospital beds and healthcare workers (*30*). In our analysis, we see a significant association between COVID-19 mortality and the rate of COVID-19 admissions. This relationship may be explained by the strain placed on hospital resources with increasing COVID-19 cases. We note that the second wave in NYC reached a lower peak number of cases with a more even distribution of admissions over the same period (*2*). In our analysis, over twice as many patients were admitted with COVID-19 during the four-month first wave compared to the four-month second wave period. This result is in line with studies associating efforts that flatten the curve of COVID-19 cases with reduced case fatality (*31*).

Lastly, differences in mortality by wave may be affected by evolution of SARS-CoV-2 and the prevalence of different viral genotypes. Wave 2 in NYC was primarily driven by multiple variants of the ancestral SARS-CoV-2 lineage (*32*). Multiple subtypes of the Iota (B.1.526) lineage were characterized in NYC during the second wave, with a high prevalence of the E484K mutation, which is associated with resistance to therapeutic monoclonal antibodies as well as convalescent and vaccinee sera (*32*). The Iota lineage was subsequently outpaced by the Alpha (B.1.1.7) variant of concern in NYC, which several studies have associated with both increased transmissibility and mortality compared to the ancestral virus (*33,34*).

### Limitations

Our study has several limitations. Cases of COVID-19 included in this analysis are likely to be undercounted from the first wave. All cases admitted with positive tests during this period would be included. Still, testing capacity was limited at the time resulting in tests being prioritized for patients with high clinical suspicion of COVID-19 or underlying comorbidities. Detection of incidental COVID-19 likely increased in the second wave when routine testing was widely available. Information bias in the EHR resulted in inadequate information to accurately characterize patients with co-morbid conditions. RT-PCR Ct data were also missing in a differential way that could have biased findings in either direction. The inclusion of patients admitted to our institution may also not wholly reflect NYC-wide cases since some individuals likely decided to avoid presentation to the hospital, especially during the first pandemic wave. In addition, it is possible that pre-existing immunity had a differential impact on infections and severe illness during the second wave. Our conclusions are limited to hospitalized patients. Extrapolating to the general population can increase the likelihood of Berkson’s bias in identifying spurious correlations not present outside the hospital setting. Our estimates of hospital capacity are based on COVID-19 admissions due to difficulties accurately estimating total hospital admissions from our database. Patients admitted with COVID-19, however, utilize specific hospital resources that would be expected to impact the care of other COVID-19 patients, including oxygen, ventilators, and ICU beds and staff. While the global population of admitted patients may not be expected to utilize the same hospital resources to the same degree, some conditions such as other respiratory viral infections, bacterial pneumonia, asthma, chronic obstructive pulmonary disease, interstitial lung disease, and heart failure may be expected to utilize similar resources and would not be accounted for in our analysis. These data would be strengthened by estimating ICU bed capacity. We reduced selection bias in our sample and model specification by right censoring patients after 30 days since the proportional odds assumption did not hold. Patients observed for longer than 30 days were a small subset and excluded from regression analyses.

### Public Health Implications

We noted a distinct reduction in COVID-19 mortality between the first and second epidemic waves in NYC associated with several covariates. The explanation for this reduction is multifactorial and likely includes standardization of COVID-19 management, availability, and knowledge of effective therapies, knowledge of ineffective treatments and interventions, as well as reduced strain on critical healthcare resources. Public health interventions are also likely to be critical contributors to the observed mortality differences given changes in lockdown policies, mask guidance, social distancing behavior, availability and speed of SARS-CoV-2 testing, and availability of vaccines for high-risk groups. A focus on the specific variables associated with reduced and increased mortality in this analysis may help prepare for future potential epidemic waves by improving the accuracy of COVID-19 projections, demographic impact, policy decisions, and public health preparations. Furthermore, plans to address future potential pandemics may benefit from prioritizing rapid, systematic methods of studying and developing treatment standards and plans to rapidly adjust hospital capacity and scale up necessary resources.

## Data Availability

All data produced in the present study are available upon reasonable request to the authors

## Supplemental material

**Table S1:**
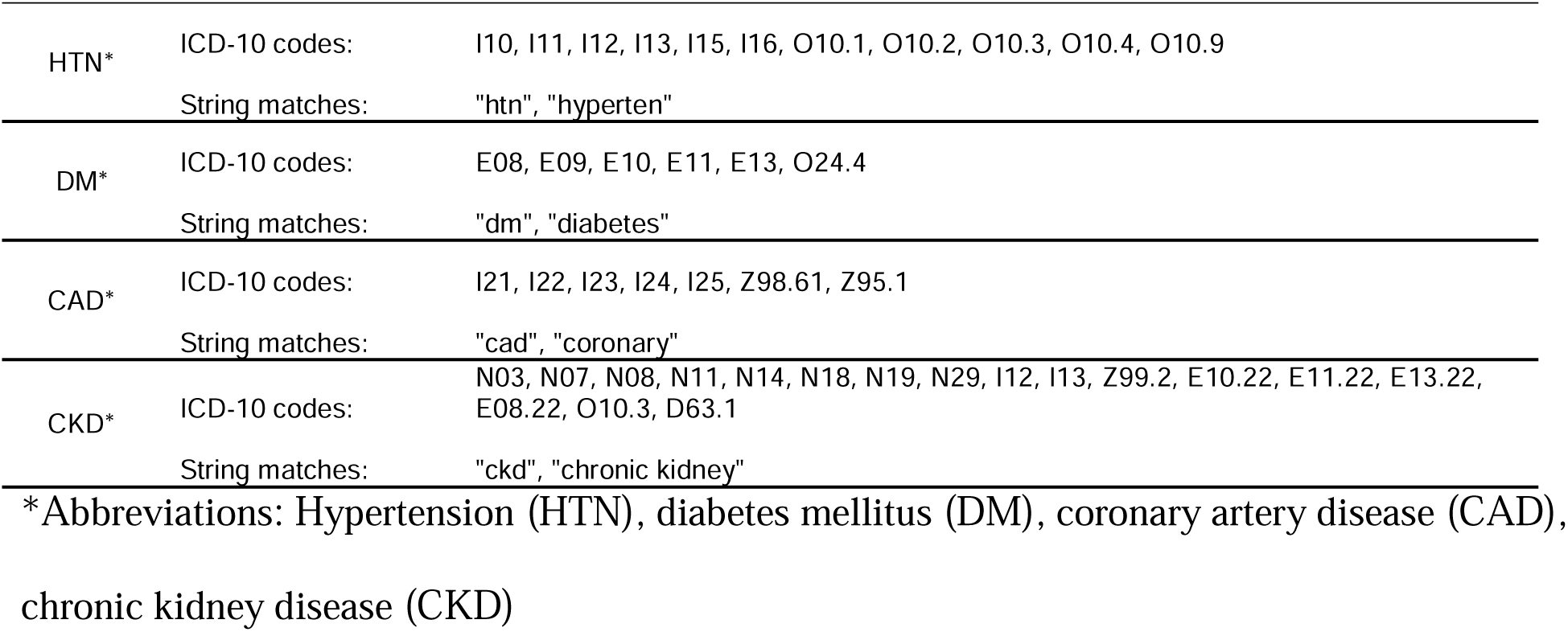
ICD-10 codes used to query patient charts to determine pre-existing conditions for each of the listed diagnoses. Electronic medical record also queried for listed text strings to determine pre-existing conditions.

|  |  |  |
| --- | --- | --- |
| HTN* | ICD-10 codes: | I10, I11, I12, I13, I15, I16, O10.1, O10.2, O10.3, O10.4, O10.9 |
|  | String matches: | "htn", "hyperten" |
| DM* | ICD-10 codes: | E08, E09, E10, E11, E13, O24.4 |
|  | String matches: | "dm", "diabetes" |
| CAD* | ICD-10 codes: | I21, I22, I23, I24, I25, Z98.61, Z95.1 |
|  | String matches: | "cad", "coronary" |
| CKD* | ICD-10 codes: | N03, N07, N08, N11, N14, N18, N19, N29, I12, I13, Z99.2, E10.22, E11.22, E13.22, E08.22, O10.3, D63.1 |
|  | String matches: | "ckd", "chronic kidney" |

\*Abbreviations: Hypertension (HTN), diabetes mellitus (DM), coronary artery disease (CAD), chronic kidney disease (CKD)

**Table S2:** Unadjusted and adjusted Cox proportional hazards model of the association between epidemic wave and death by 30 days after admission adjusted for potentially confounding factors, among those ≥ 75 years old.

|  | Unadjusted<br>HR (95% CIs) | Adjusted for epi-period |  |
| --- | --- | --- | --- |
|  |  | HR for covariate (95% CIs)<br>adjusted for Epidemic<br>period | HR for epidemic period<br>(95% CIs) adjusted each<br>covariate individually |
| Epidemic Period |  |  |  |
| First | Ref | NA | Ref |
| Second | **0.43 (0.35, 0.53)** |  | **0.43 (0.35, 0.53)** |
| Sex |  |  |  |
| Female | Ref | Ref | Ref |
| Male | 1.14 (0.95, 1.36) | 1.11 (0.93, 1.33) | **0.43 (0.35, 0.54)** |
| Race/Ethnicity |  |  |  |
| Hispanic/Latino | Ref | Ref | Ref |
| Non-Hispanic Black | 1.14 (0.84, 1.54) | 1.10 (0.81, 1.49) | **0.43 (0.35, 0.53)** |
| Non-Hispanic White | 1.08 (0.83, 1.40) | 1.08 (0.84, 1.41) |  |
| Other | 0.97 (0.77, 1.22) | 0.98 (0.78, 1.23) |  |
| BMI (kg/m2) |  |  |  |
| <30 | Ref | Ref | Ref |
| $\geq 30$ | 1.23 (0.99, 1.53) | 1.20 (0.96, 1.50) | **0.48 (0.38, 0.59)** |
| Ct value, median (IQR) | **0.98 (0.97, 0.99)** | **0.98 (0.96, 0.99)** | **0.35 (0.22, 0.55)** |
| Viral load categories |  |  |  |
| Low (Ct >32 or 30) | Ref | Ref | Ref |
| Medium (Ct 27-32 or 25-30) | **1.61 (1.22, 2.12)** | **1.62 (1.23, 2.14)** | **0.35 (0.22, 0.56)** |
| High (Ct <27 or 25) | **1.72 (1.35, 2.20)** | **1.77 (1.39, 2.26)** |  |
| Ever DNI |  |  |  |
| Yes | **6.81 (5.11, 9.06)** | **6.22 (4.66, 8.29)** | Ref |
| No | Ref | Ref | **0.54 (0.44, 0.68)** |
| Oxygen level at presentation |  |  |  |
| Room Air | Ref | Ref | Ref |
| Nasal Cannula | **2.61 (1.93, 3.54)** | **2.55 (1.88, 3.46)** | **0.66 (0.52, 0.82)** |
| Non-rebreather | **8.65 (6.44, 11.6)** | **7.58 (5.60, 10.25)** |  |
| Non-invasive ventilation | **4.79 (2.44, 9.39)** | **4.78 (2.44, 9.37)** |  |
| Intubation | **7.82 (5.18, 11.9)** | **6.97 (4.60, 10.57)** |  |
| ICU admission by time, N (%) |  |  |  |
| Non-ICU | Ref | Ref | Ref |
| ICU at presentation | **2.54 (1.97, 3.27)** | **2.36 (1.83, 3.04)** | **0.44 (0.35, 0.54)** |
| ICU after presentation | **2.02 (1.51, 2.71)** | **2.14 (1.60, 2.86)** |  |
| Steroid use, N (%) |  |  |  |
| No | Ref | Ref | Ref |
| Yes | 1.00 (0.83, 1.20) | **1.49 (1.22, 1.83)** | **0.37 (0.29, 0.47)** |
| Remdesivir use, N (%) |  |  |  |
| No | Ref | Ref | Ref |
| Yes | **0.76 (0.58, 0.98)** | **1.69 (1.20, 2.37)** | **0.34 (0.26, 0.45)** |
| Hospital admission volume divided by<br>50 (per week) | **1.10 (1.08, 1.12)** | **1.08 (1.05, 1.11)** | **0.72 (0.53, 0.97)** |

**Table S3:** Cox proportional hazards model of the association between steroid use and 30-day mortality stratified to examine confounding by disease severity.

| Stratifying variable | Treatment Variable | Dead N | Alive N | HR (95% CI) |
| --- | --- | --- | --- | --- |
|  | Steroid<sup>+</sup> | 338 | 1384 | 1.15 (1.00, 1.33) |
|  | No Steroid | 405 | 2052 |  |
| Age < 75 years | Steroid<sup>+</sup> | 149 | 1034 | 1.47 (1.17, 1.85) |
|  | No Steroid | 139 | 1491 |  |
| Age ≥ 75 years | Steroid<sup>+</sup> | 189 | 350 | 1.00 (0.92, 1.20) |
|  | No Steroid | 266 | 561 |  |
| Wave 1 | Steroid<sup>+</sup> | 185 | 527 | **1.34 (1.12, 1.59)** |
|  | No Steroid | 390 | 1668 |  |
| Wave 2 | Steroid<sup>+</sup> | 153 | 857 | **4.2 (2.51, 7.26)** |
|  | No Steroid | 15 | 384 |  |
| Wave 1, ICU | Steroid<sup>+</sup> | 94 | 202 | **0.53 (0.41, 0.70)** |
|  | No Steroid | 130 | 144 |  |
| Wave 1, Non-ICU | Steroid<sup>+</sup> | 91 | 325 | 2.22 (0.97, 5.09) |
|  | No Steroid | 260 | 1524 |  |
| Wave 2, ICU | Steroid<sup>+</sup> | 79 | 107 | **1.48 (1.17, 1.88)** |
|  | No Steroid | 6 | 22 |  |
| Wave 2, Non-ICU | Steroid<sup>+</sup> | 74 | 750 | **3.83 (1.92, 7.64)** |
|  | No Steroid | 9 | 362 |  |
|  | Remdesivir<sup>+</sup> | 108 | 575 | **0.81 (0.66, 0.99)** |
|  | No Remdesivir | 660 | 2922 |  |
| Age < 75 years | Remdesivir<sup>+</sup> | 42 | 420 | 0.85 (0.62, 1.17) |
|  | No Remdesivir | 251 | 2157 |  |
| Age ≥ 75 years | Remdesivir<sup>+</sup> | 66 | 155 | **0.76 (0.58, 0.98)** |
|  | No Remdesivir | 409 | 765 |  |
| Wave 1 | Remdesivir<sup>+</sup> | 8 | 74 | **0.42 (0.21, 0.84)** |
|  | No Remdesivir | 592 | 2172 |  |
| Wave 2 | Remdesivir<sup>+</sup> | 100 | 501 | **2.06 (1.51, 2.80)** |
|  | No Remdesivir | 68 | 750 |  |
| Wave 1, ICU | Remdesivir<sup>+</sup> | 6 | 34 | **0.30 (0.13, 0.67)** |
|  | No Remdesivir | 225 | 319 |  |
| Wave 1, Non-ICU | Remdesivir<sup>+</sup> | 2 | 40 | 0.27 (0.07, 1.07) |
|  | No Remdesivir | 367 | 1853 |  |
| Wave 2, ICU | Remdesivir<sup>+</sup> | 45 | 68 | 0.95 (0.62, 1.45) |
|  | No Remdesivir | 40 | 61 |  |
| Wave 2, Non-ICU | Remdesivir<sup>+</sup> | 55 | 433 | **2.96 (1.88, 4.67)** |
|  | No Remdesivir | 28 | 689 |  |

**Figure S1.**
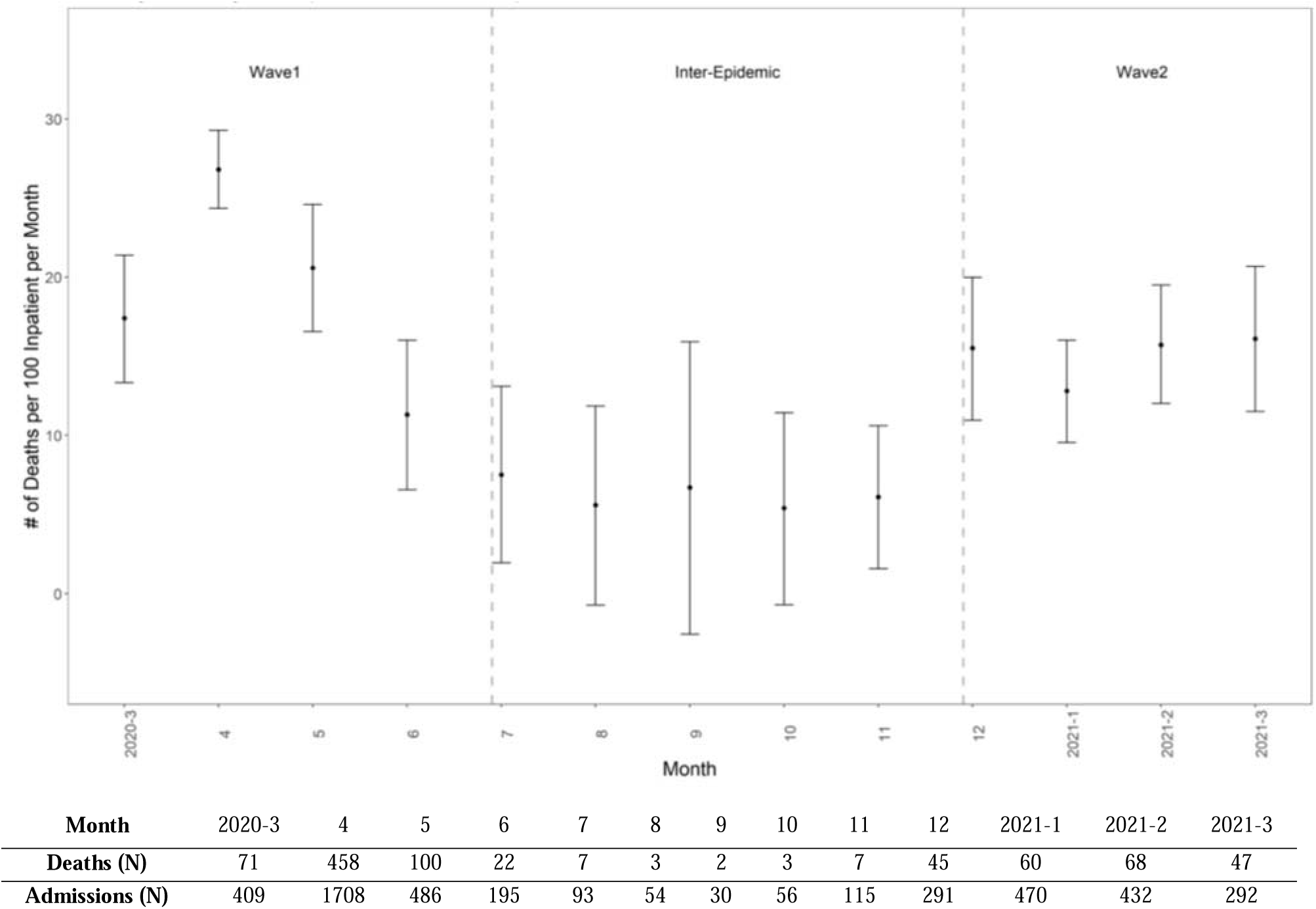
In-hospital monthly mortality rates per 100 patients and 95% confidence bounds among 4132 patients admitted to the medical center between March 2020 and March 2021.

